# Estimating Cuff-less Continuous Blood Pressure from Fingertip Photoplethysmogram Signals with Deep Neural Network Model

**DOI:** 10.1101/2022.03.14.22272354

**Authors:** Yu Chen

## Abstract

**Objective:** Blood pressure (BP) is an important physiological index reflecting cardiovascular function. Continuous blood pressure monitoring helps to reduce the prevalence and mortality of cardiovascular diseases. In this study, we aim to estimate systolic blood pressure (SBP) and diastolic blood pressure (DBP) values continuously based on fingertip photoplethysmogram (PPG) waveforms using deep neural network models.

**Methods:** Two models were proposed and both models consisted of three stages. The only difference between them was the method of extracting features from PPG signals in the first stage. Model 1 adopted Bidirectional Long Short-Term Memory (BiLSTM), while the other used convolutional neural network. Then, the residual connection was applied to multiple stacked LSTM layers in the second stage, following by the third stage with two fully connected layers.

**Results:** Our proposed models outperformed other methods based on similar dataset or framework, while in our proposed models, the model 2 was superior to model 1. It satisfied the standard of Association for the Advancement of the Medical Instrumentation (AAMI) and obtained grade A for SBP and DBP estimation according to the British Hypertension Society (BHS) standard. The mean error (ME) and standard deviation (STD) for SBP and DBP estimations were 0.21 ± 6.40 mmHg and 0.19 ±4.71 mmHg, respectively.

**Conclusion:** Our proposed models could extract important features of fingertip PPG waveforms automatically and realize cuff-less continuous BP monitoring, which can be helpful in the identification and early treatment of abnormal blood pressure, thus may reduce the occurrence of cardiovascular malignant events.

## 1. Introduction

Blood pressure (BP) is the pressure of blood acting on the walls of arteries[1], which fluctuates regularly with the contraction and relaxation of the heart. The maximum pressure corresponding to the contraction of the heart is called systolic blood pressure (SBP), and the minimum pressure corresponding to the diastole is called diastolic blood pressure (DBP), the normal and high-normal ranges of SBP and DBP in adults are <140 mmHg and <90 mmHg, respectively [2, 3]. High SBP is one of the main risk factors of cardiovascular diseases (CVDs), especially ischemic heart disease (IHD) and stroke, which are the leading cause of global mortality and disability[4]. With the acceleration of population aging, the prevalence of adults with high SBP nearly doubled from 2.18 billion in 1990 to 4.06 billion in 2019[4]. From 1990 to 2019, the number of deaths due to high SBP increased from 6.79 million to 10.8 million[4]. Therefore, BP monitoring is critical for the prevention and treatment of CVDs, alleviating the disease burden, and realizing the global health.

The existing methods of BP measurement can be divided into invasive measurement and non-invasive measurement. The invasive method usually places a catheter with a BP sensor directly in the blood vessel or heart for measurement. This method is the most accurate and considered as the gold standard of BP measurement internationally. However, it has the risk of bleeding and infection, which is only suitable for critically ill patients in hospitals and should be operated by professionals[5, 6]. Compared with invasive method, non-invasive measurement is more acceptable by people. Non-invasive BP measurement includes intermittent blood pressure measurement (IBPM) and continuous blood pressure measurement (CBPM). The two traditional IBPM methods commonly used in clinical practice are the manual method based on Korotkoff sounds and the automatic method based on cuff pressure oscillation[7, 8]. Both methods are susceptible to external interference and require pressure on the cuff, which will cause discomfort to the human body. Besides, by the influence of genetic, environmental, and lifestyle factors, BP is dynamic[1]. For patients who need to monitor BP closely, in contrast to IBPM, CBPM can provide more detailed information of BP changes, which is of great significance in the diagnosis and analysis of diseases and medical research. The CBPM methods mainly include the volume-compensation, arterial tonometry, and pulse wave measurement[9]. The first two methods can also cause discomfort for its pressure on the blood vessels[10, 11]. In addition, the measurement process of the volume-compensation method is complex, and any state that causing low peripheral perfusion will affect the measurement results, such as cold temperature, vascular disease, and Raynaud’s disease[9, 10]. Moreover, the arterial tonometry method has a high requirement for sensor positioning, so it is rarely used in clinical practice[9, 11].

In recent years, pulse wave measurement has become one of the most promising methods for non-invasive cuff-less CBPM, especially the method based on photoplethysmogram (PPG)[6]. PPG is a non-invasive optical measurement technology for measuring peripheral pulse blood volume changes, which can be obtained from ear, finger, toe, and other sites[12]. It has been widely used in clinical physical monitoring, vascular assessment, and automatic function, with the characteristics of simple, reliable, and low cost[12]. The summary of the BP measurement methods is shown in figure 1.

**Figure 1.**
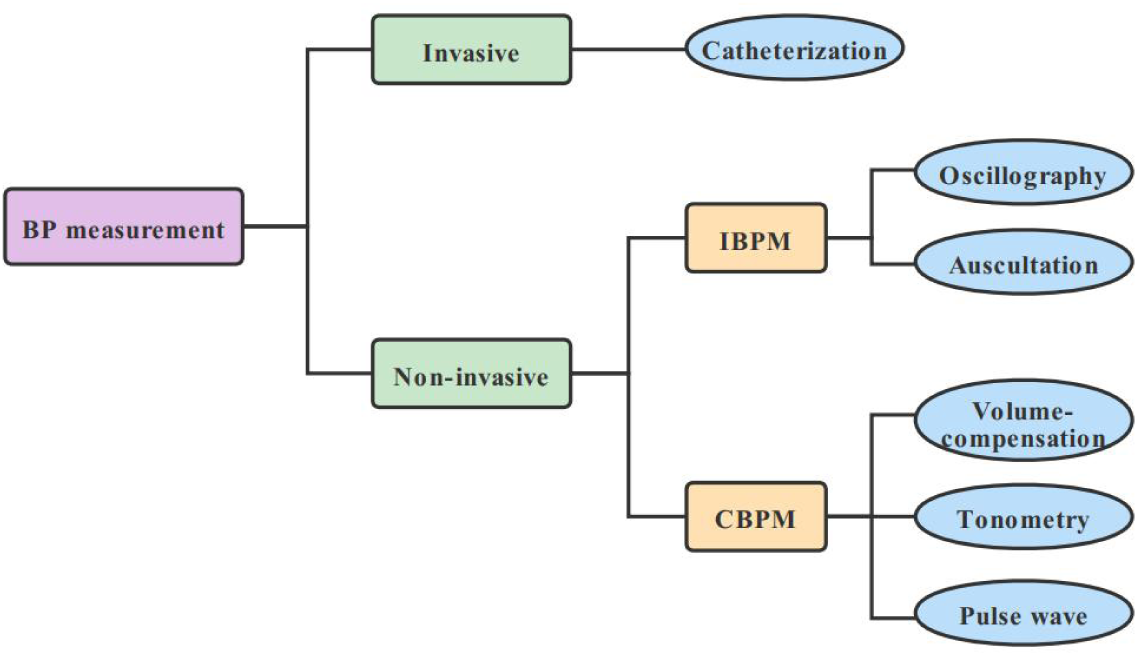
The block diagram of classification for BP measurement methods. BP = blood pressure, IBPM = intermittent blood pressure measurement, CBPM = continuous blood pressure measurement.

As a typical diagnosis and treatment method of traditional Chinese medicine (TCM), the pulse diagnosis has a history of more than two thousand years. It’s one of the important means for TCM practitioners to obtain disease information[13]. TCM practitioners place their finger pulps on the skin of the human body where the pulsation is obvious, such as Renying (the carotid artery), Cunkou (the radial artery), and Fuyang (the dorsalis pedis artery) to obtain disease information[14, 15]. Considering the convenience of pulse-taking, Cunkou is usually chosen. However, the sensitivity of finger pulps is limited, it can only feel the pulse information from somewhere with obvious pulsation[15]. Besides, the pulse diagnosis relies on the subjective practical experience of the clinicians, which is difficult to be standardized and popularized in clinical practice. In recent years, with the rapid development of modern technology, many new methods for obtaining or analyzing pulse information have emerged, which bring new opportunities for the objective research of TCM[13]. For instance, the aforementioned PPG method can sensitively measure pulse information at many sites of the human body, other than Renying, Cunkou, and Fuyang.

The aim of this study is to explore the relationship between pulse and blood pressure based on fingertip PPG signals using neural network model, and to obtain continuous blood pressure information from the pulse wave. The establishment of the present method may also contribute our understanding of the relation between pulse and health in TCM theory. The present paper is organized as follows: Section 2 introduces the related methods and models of BP estimation and describes the proposed method briefly. Section 3 give details about the data source and processing as well as the overall framework. Section 4 reports our experimental results. Section 5 discusses the results. Finally, section 6 concludes the paper.

## 2. Background

### 2.1 Morphological characteristics of PPG signal

PPG waveform consists of two parts: a pulsatile physiological waveform (‘AC’ component) and a slowly varying baseline (‘DC’ component). The rich information of heart pulsation mainly exists in the ‘AC’ component of the PPG waveform, which is mainly composed of the anacrotic phase and the catacrotic phase[12]. The rising border of the pulse corresponds to the anacrotic phase and the falling border represents the catacrotic phase (Figure 2). These two phases correspond to the systole and diastole of the heart respectively. During the relaxation of the heart, due to the wave reflections from the periphery, a dicrotic notch is formed in the catacrotic phase, which is commonly seen in healthy people[6, 12].

**Figure 2.**
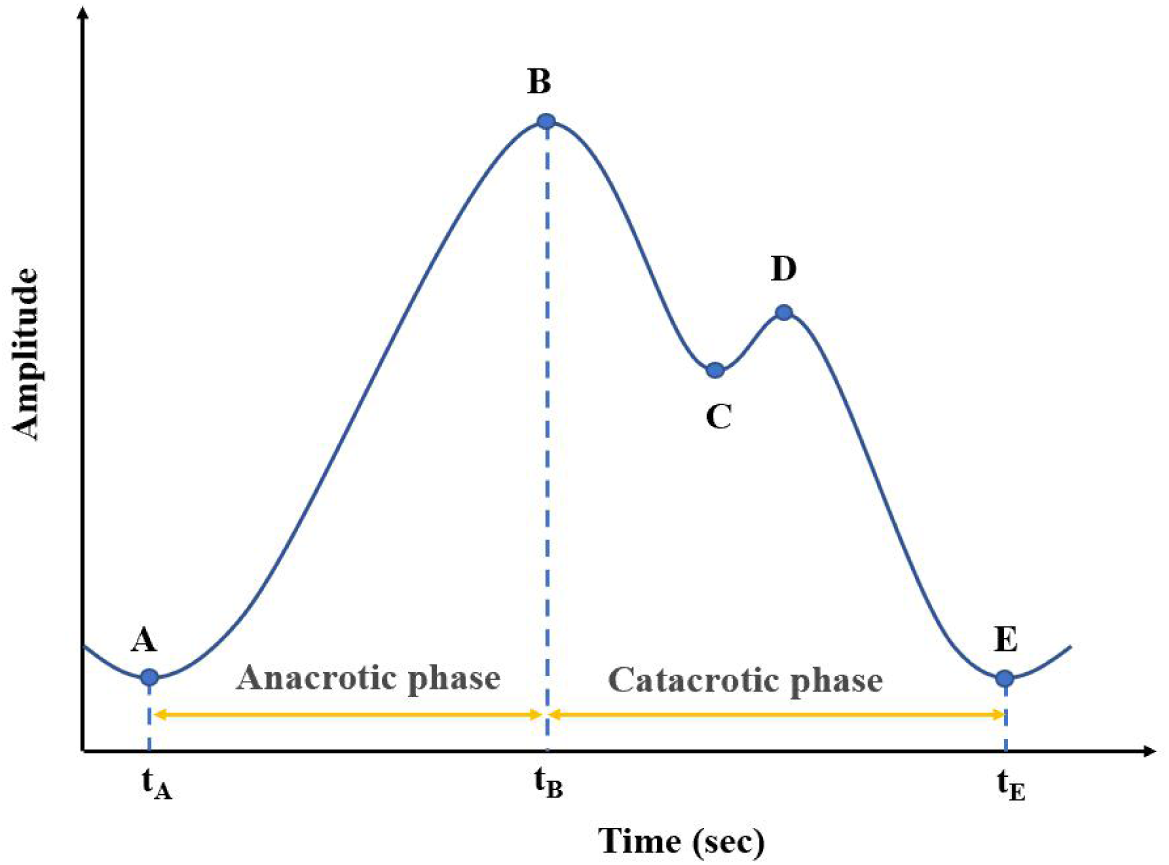
The sketch map of the ‘AC’ component of typical PPG waveform. A refers to the starting point of this wave, B is the main peak, C corresponds to the dicrotic notch, D represents the dicrotic wave, and E is the starting point of the next wave.

### 2.2 BP estimation based on PPG

The potential of PPG to measure BP has been recognized for decades[12]. In this section, we described several methods of BP measurement based on PPG signals.

#### 2.2.1 Pulse transit time methods

Pulse transit time (PTT) is usually defined as the travel time between aortic valve opening and arrival of the blood flow to the distal position[2]. In many literatures, PTT is usually replaced by pulse arrival time (PAT) for its starting point is difficult to confirm[2, 16, 17]. PAT refers to the time difference between the R-peak of an electrocardiogram (ECG) and a specific feature point of a PPG[18]. For example, Fung, et al. [19] have evaluated the BP value from the PTT (PAT actually), which can be described by Equation (1).

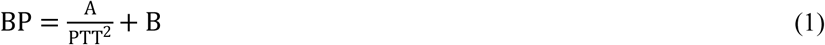

A and B are constants associated with subjects. This PTT-BP algorithm is simplistic, but it requires two sensors and complex calibration, which is hard to be applied in practice[19].

#### 2.2.2 Pulse wave velocity methods

Pulse wave velocity (PWV) is the velocity of the pressure wave propagation in the blood vessels[5], which mainly depends on the elastic and geometric properties of the arterial wall[20]. The relationship of them can be illustrated using Moens-Kortweg equation[21, 22]. Besides, PWV can be calculated by dividing the distance L between two sensors on the same arterial branch by the time difference T for the pressure wave propagating at this distance[23]. Finally, the calculation of BP values from PWV can be described by Equation (2).

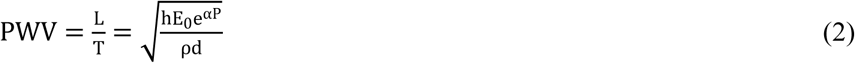

where h is the thickness of the vessel wall, E0 represents the Young’s modulus of elasticity for zero arterial pressure, e is Euler’s number (approximately 2.718), α is a vessel parameter (typically 0.016 mmHg^-1^ to 0.018 mmHg^-1^), P refers to BP, ρ represents blood density and d indicates the diameter of the vessel. Since parameters such as the arterial elasticity and the length of the artery vary between individuals, it needs frequent calibration. Hence, it affects the application of PWV in health care[6].

#### 2.2.3 Pulse wave analysis methods

Pulse wave analysis (PWA) refers to extract important features from the PPG waveform directly for creating models using machine learning or deep neural network[6, 24]. The most significant advantage of this method is that it only requires one PPG sensor for BP estimation[6]. Currently, there are two main methods for feature extraction. One is manual feature extraction, which is complex, error prone and uncompleted. For instance, Lin, et al. [25] have extracted 65 features from the PPG signals and their first and second derivative values for BP estimation and indicated that their proposed features set outperforms the two previously proposed feature sets[26, 27]. The other is automatic feature extraction based on specific algorithm[17]. Esmaelpoor, et al. [5] have adopted convolutional neural network (CNN) to extract the morphological features from PPG waveforms and then transmitted the extracted feature vector to the next stage for BP values evaluation. As the PPG signals can be affected by kinds of internal and external factors, extracting features manually become extraordinarily difficult[17]. Therefore, automatic feature extraction has more advantages for its simplicity and convenience.

### 2.3 Machine learning models

In recent years, many linear and non-linear models have been employed to estimate BP with PPG signals, such as linear regression[28, 29], support vector machine (SVM)[30], AdaBoost classifier[31], feedforward neural network[32], restricted boltzmann machine[33], and recurrent neural network (RNN)[34]. Generally, the performance of nonlinear model is better than that of linear model, but it also depends on the quality of dataset and the modelling method [6]. Currently, some advanced approaches have been proposed, such as Long-Short Term Memory (LSTM)[2, 17] and CNN models[5]. These models perform well in predicting BP values by using automatically extracted important features and have great potential in the future.

In this work, we proposed two hybrid neural network models to estimate cuff-less real-time BP values utilizing the features automatically extracted from fingertip PPG signals. The two hybrid models were similar in structure and mainly consisted of three stages. The first stage utilized CNN or Bidirectional Long Short-Term Memory (BiLSTM) to extract important features automatically from fingertip PPG signals, and the remaining stages used stacked LSTM layers with residual connection and two fully connected (FC) layers to estimate BP values based on the features extracted in the first stage.

## 3. Materials and methods

### 3.1 Data source

The dataset used in this paper was derived from [35], which belongs to the Multi-parameter Intelligent Monitoring in Intensive Care (MIMIC) II online waveform database of PhysioNet[36]. This database provides rich data including PPG (collected from pulse oximeter placed on the fingertip) as well as the arterial blood pressure (ABP, measured by a catheter in the radial artery) with sampling frequency of 125Hz. While the database provides a wealth of raw waveform signals, their quality is uneven, therefore, a powerful preprocessing method was taken by [35] for removing undesirable components such as baseline wander and low-quality signals to obtain reliable records. Thus, the preprocessed dataset was applied to evaluate the performance of our methods proposed in this work.

### 3.2 Preprocessing

#### 3.2.1 Waveform segmentation and abnormal BP signals processing

The waveforms of PPG and ABP always fluctuate periodically with the heartbeat, however, the fluctuation frequency is dynamic by the influence of internal and external factors. Therefore, the method of sliding segmentation with fixed number of cycles was adopted in this work rather than a fixed length. As shown in Figure 3, each segment consists of two cycles with the length of three consecutive PPG peaks, and then extracts the corresponding ABP segments according to the peaks of PPG. It’s worth noting that there is an overlap between adjacent segments with a length of one cycle.

**Figure 3.**
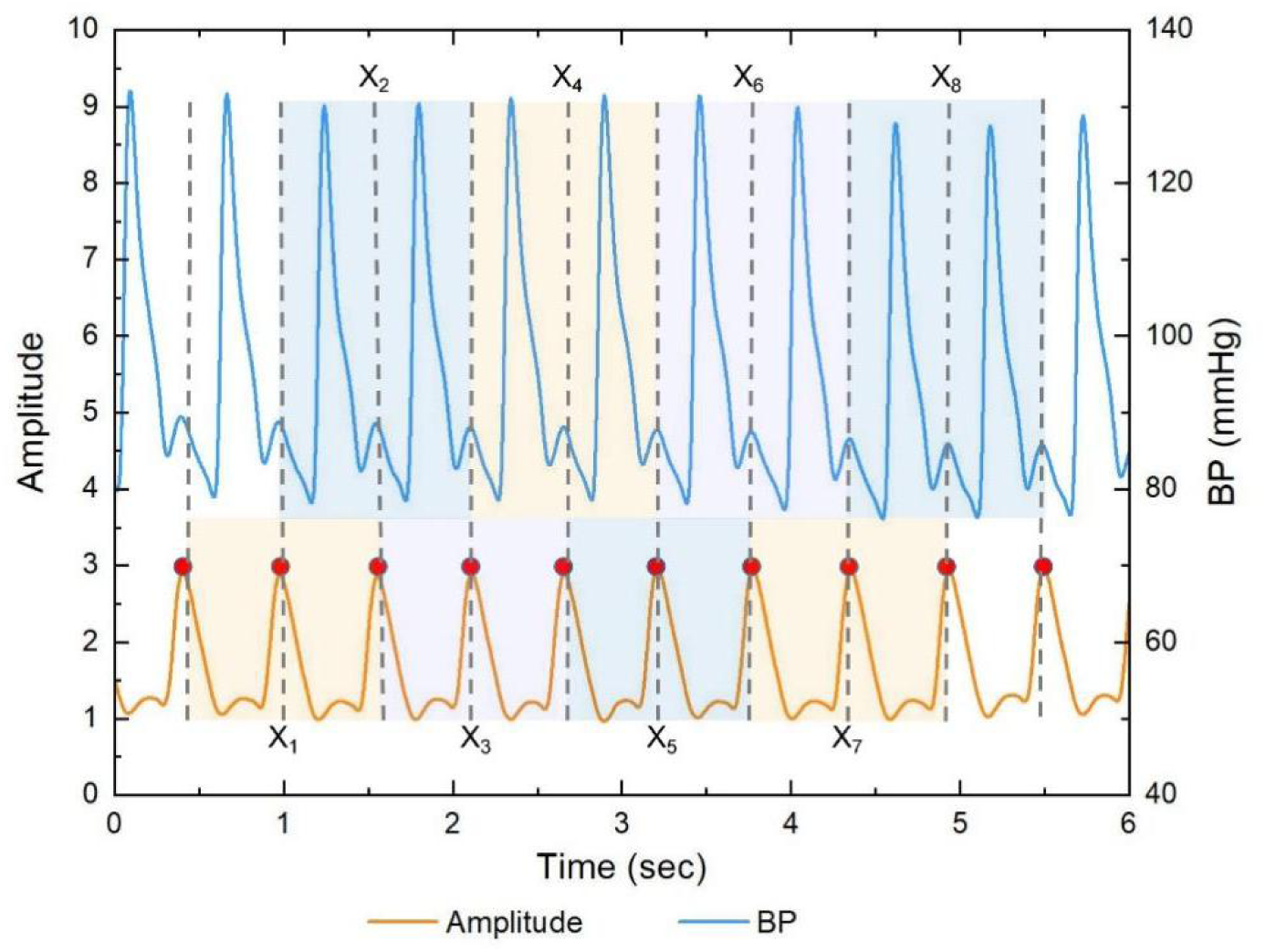
Illustration of waveform segmentation. The waveform of PPG is showed by orange, the ABP signal is performed by blue, and the peaks of PPG are marked with red dots. BP = blood pressure; ABP = arterial blood pressure; PPG = photoplethysmogram.

After removing the ABP segments with very high or very low BP values (e.g., SBP ≥ 180, DBP ≥ 130, SBP ≤ 80, DBP ≤ 60) and the corresponding segments of PPG (SBP and DBP correspond to the maximum and minimum of each ABP segment respectively), we obtained millions of PPG segments and then resampled them to 256 samples. Finally, the first 100,000 segments of the dataset were selected in this paper considering the training cost of models. Table 1 and figure 4 show the distribution and ranges of SBP, DBP, and mean arterial pressure (MAP) values in the final dataset.

**Table 1.** Statistics of the BP datasets used in the experiments.

|  | Min(mmHg) | Max(mmHg) | Mean(mmHg) | STD(mmHg) |
| --- | --- | --- | --- | --- |
| **SBP** | 83.00 | 180.00 | 134.76 | 14.28 |
| **DBP** | 60.00 | 130.00 | 70.79 | 8.84 |
| **MAP** | 68.00 | 142.00 | 92.12 | 8.14 |

**Figure 4.**
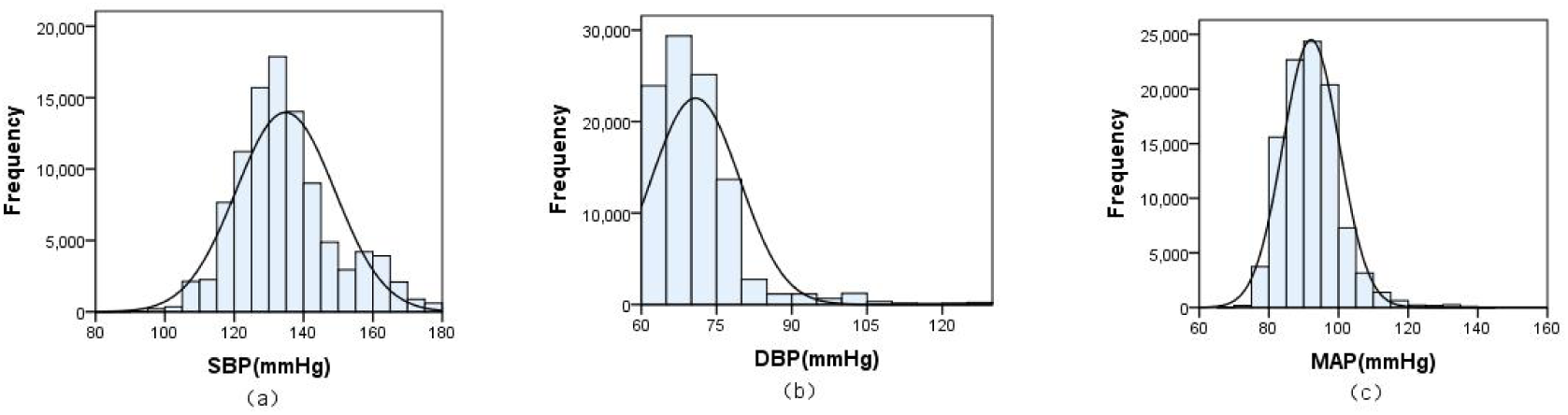
Distribution histogram of the dataset used in this paper. (a) systolic blood pressure (SBP), (b) diastolic blood pressure (DBP), (c) mean arterial pressure (MAP).

#### 3.2.2 Normalization

In order to speed up the optimization process, we normalized the amplitude of PPG signals to [0-1]. The formula is as follows:

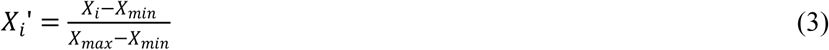

*X*_*max*_ and *X*_*min*_ are the maximum and minimum values of the training set respectively.

### 3.3 Multistage model

The overall flow diagram of the proposed methods is exhibited in figure 5. We fed the preprocessed data into two multistage deep learning models separately to test their performances in predicting BP, and the detailed frameworks are displayed in figure 6 and 7. Both models were composed of three stages. The only difference between them was in the first stage: model 1 adopted BiLSTM but model 2 adopted CNN to fully extract the important features of the whole input sequences. To solve the problem of vanishing or exploding gradient, we applied residual connections in multiple stacked LSTM layers in the next phase. Finally, the estimated SBP and DBP values were outputted after the last two fully connected (FC) layers.

**Figure 5.**
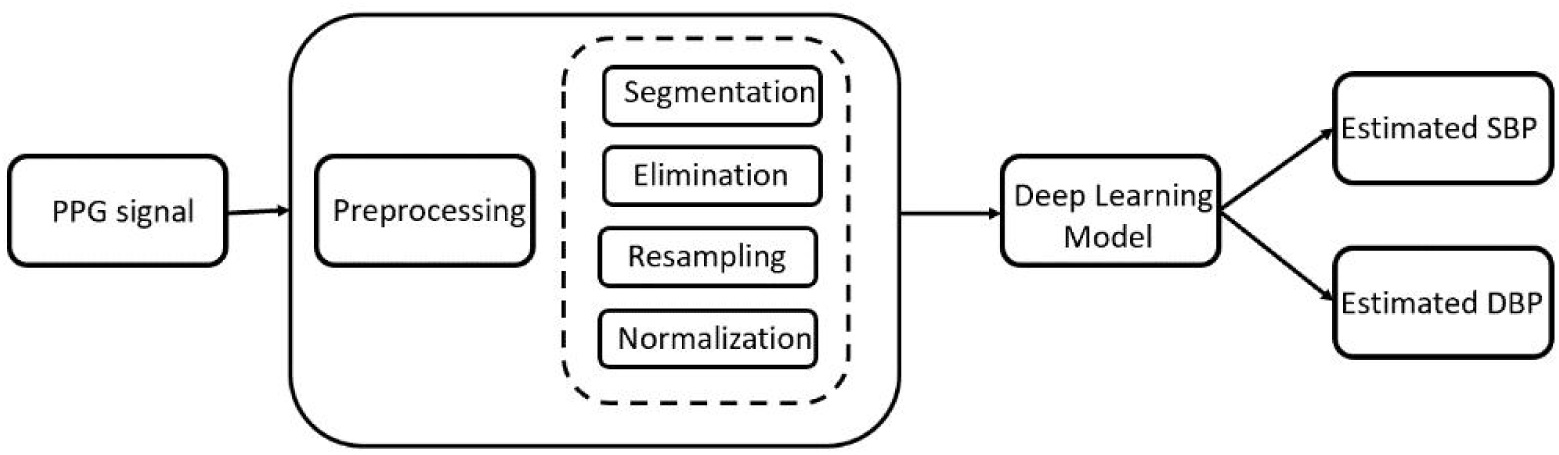
The flow diagram of proposed methods.

**Figure 6.**
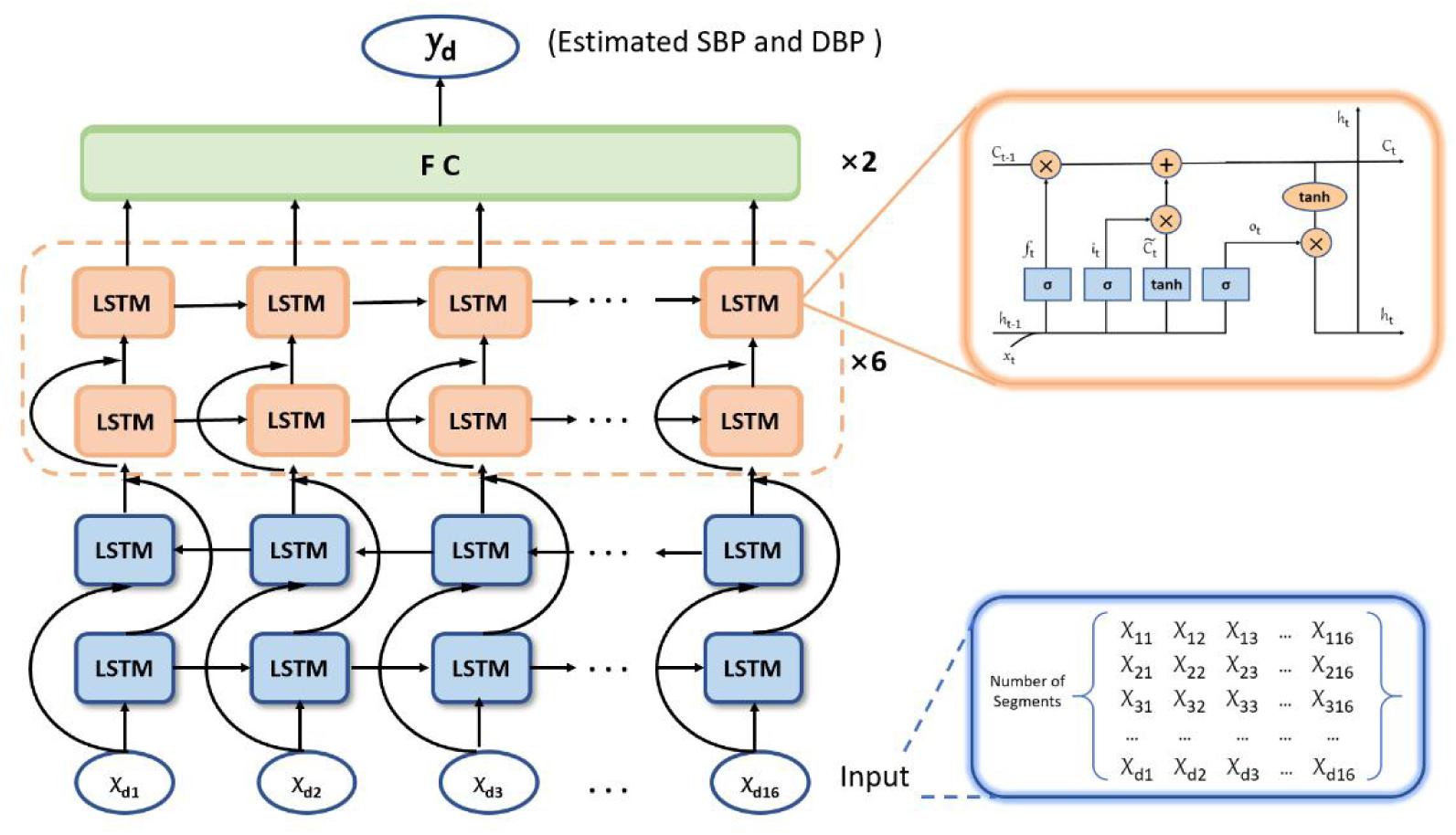
Overall framework of model 1 proposed in this paper. FC = fully connected layer, LSTM = Long-Short Term Memory.

**Figure 7.**
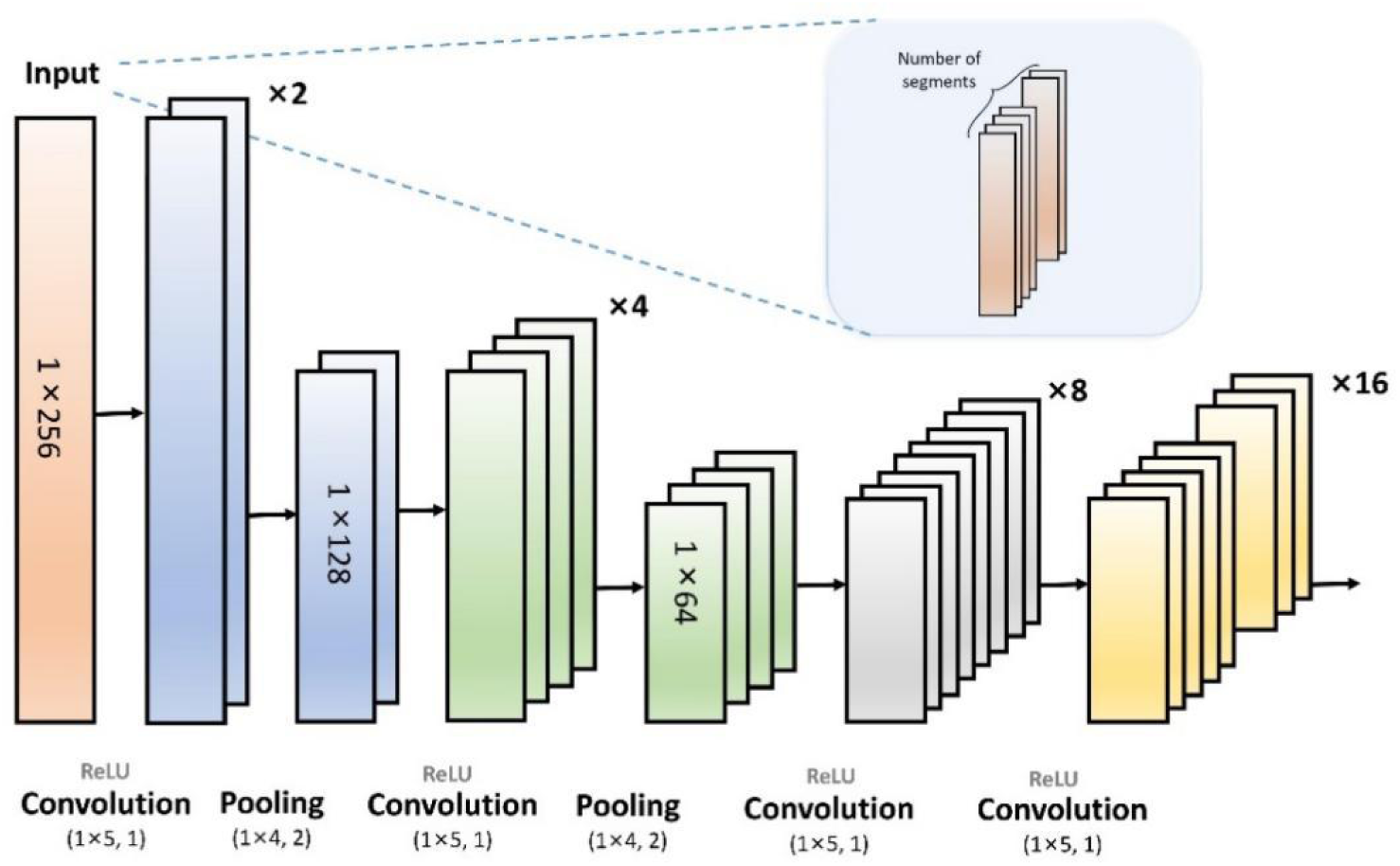
Illustration of the feature vectors extraction part in model 2.

The first stage of model 1 was a BiLSTM, which processed sequences of inputs from the front and back directions[37]. It can improve the performance of predicting when processing time series compared with unidirectional LSTM[38, 39]. In addition, CNNs have also been widely used in BP estimation and achieved good results for its advantages in local perception and parameter sharing so as to simplify the complexity of the network model[40, 41]. CNNs have been commonly used for image processing, here we adopted it to process one-dimensional time series data which were arranged in an instant sequence of time[42]. As a note, both LSTM layers of BiLSTM consisted of 32 units, each vector X in model 1 contained 16 features, and d represented the number of segments. In model 2, the CNN was composed of four hidden convolutional layers and the first two layers followed with an average pooling layer separately (please see Figure 7 for more details).

The second stage was composed of multiple stacked LSTM layers and each layer consisted of 64 units. Every two layers of LSTM were regarded as a whole, in which the residual connection was applied to the first layer. The sum of input and output vectors of previous layer was conducted as the input of next layer. In total, there were six structure cells like this (Figure 6).

In the final stage, the output from the last LSTM layer of the second stage was fed into two FC layers to predict SBP and DBP values. The number of neurons in the first FC layer was set to 512 and the second layer was 2.

Model parameters of the two models were identical. The activation function used in our proposed models was ReLU, and the dropout rate was set to 0.2. In training process, we used Adam Optimizer with the initial learning rate of 0.0001. In addition, the L2 norm of regularization was set to 0.05, and the batch size was 32. For maximal epochs, 300 was used. We used mean square error (MSE) loss function to optimize gradient.

## 4. Results

In order to obtain a more stable and reliable model, we conducted the five-fold cross-validation on the dataset using the above two models respectively and the results were displayed in Table 2 and 3. Here, we used mean absolute error (MAE) and standard deviation (STD) as the evaluation metrics of the models. We found that the performances of both models were relatively stable. In general, both algorithms we proposed achieved relatively satisfactory results, while the performance of the model 2 based on CNN is slightly better than model 1 based on BiLSTM in stability and accuracy.

**Table 2.**
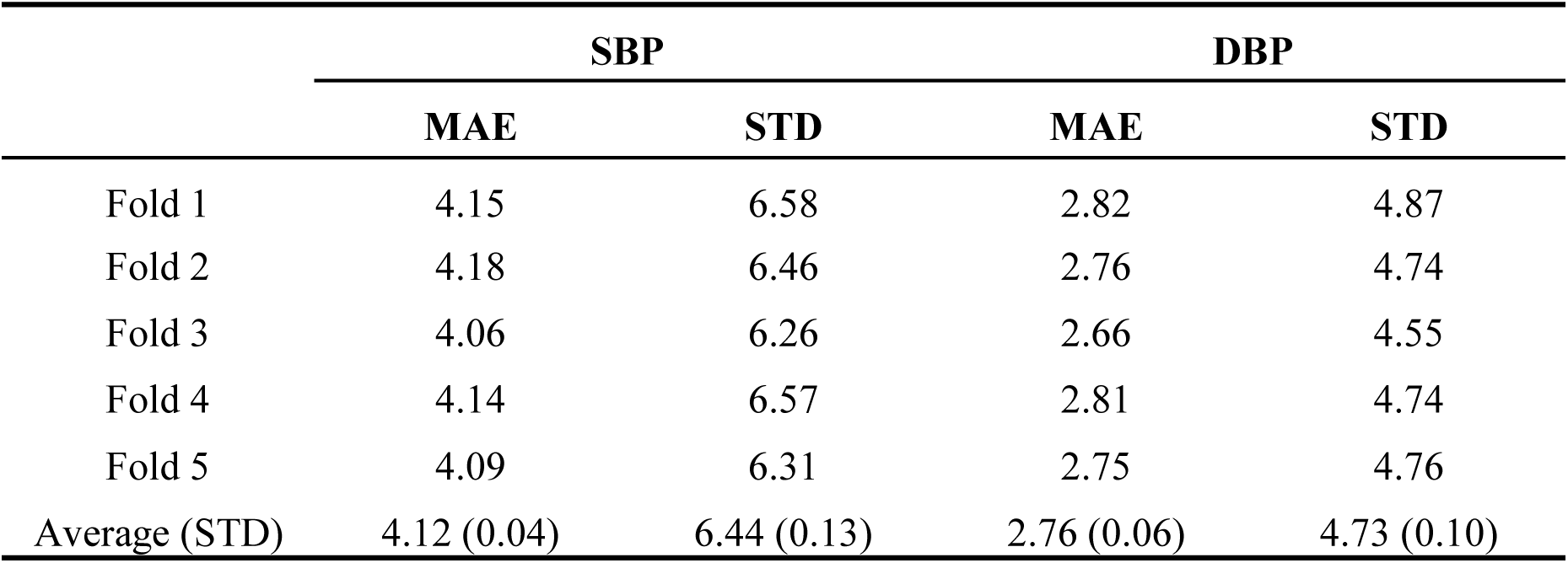
Five-fold cross-validation results of Model 1 (BiLSTM).

|  | SBP |  | DBP |  |
| --- | --- | --- | --- | --- |
|  | MAE | STD | MAE | STD |
| Fold 1 | 4.15 | 6.58 | 2.82 | 4.87 |
| Fold 2 | 4.18 | 6.46 | 2.76 | 4.74 |
| Fold 3 | 4.06 | 6.26 | 2.66 | 4.55 |
| Fold 4 | 4.14 | 6.57 | 2.81 | 4.74 |
| Fold 5 | 4.09 | 6.31 | 2.75 | 4.76 |
| Average (STD) | 4.12 (0.04) | 6.44 (0.13) | 2.76 (0.06) | 4.73 (0.10) |

**Table 3.** Five-fold cross-validation results of Model 2 (CNN-LSTM).

|  | SBP |  | DBP |  |
| --- | --- | --- | --- | --- |
|  | MAE | STD | MAE | STD |
| Fold 1 | 3.89 | 6.31 | 2.60 | 4.68 |
| Fold 2 | 3.95 | 6.34 | 2.63 | 4.68 |
| Fold 3 | 3.98 | 6.29 | 2.64 | 4.59 |
| Fold 4 | 4.05 | 6.57 | 2.70 | 4.74 |
| Fold 5 | 4.06 | 6.48 | 2.70 | 4.85 |
| Average (STD) | 3.99 (0.06) | 6.40 (0.11) | 2.65 (0.04) | 4.71 (0.09) |

The comparation of our proposed models with other algorithms in BP estimation were shown in Table 4. Notably, the estimation error (indicated by MAE and STD) of SBP and DBP of our proposed models were smaller than that of other works. Besides, our proposed models did not require manual feature extraction, which avoids the impact of manual operation on the training process. Obviously, we found that the DBP estimation accuracy of all models in Table 4 were higher than that of SBP, which may be caused by the large fluctuation of SBP in human bodies. Therefore, the estimation error of SBP should be the focus of evaluating the performance of models in predicting BP.

**Table 4.** Performance of different algorithms in blood pressure estimation.

| Methods | Dataset | Sensors | SBP(mmHg) |  | DBP(mmHg) |  |
| --- | --- | --- | --- | --- | --- | --- |
|  |  |  | MAE | STD | MAE | STD |
| MARS[43] | MIMIC II | PPG&ECG | 7.83 | 9.1 | 4.86 | 5.21 |
| AdaBoost[26] | MIMIC II | PPG&ECG | **11.17** | 10.09 | **5.35** | 6.14 |
| BiLSTM-4L[2] | MIMIC II | PPG&ECG | 6.73 | **14.51** | **2.52** | **6.44** |
| Model 1 (BiLSTM) | MIMIC II | PPG | 4.12 | 6.44 | 2.76 | 4.73 |
| Model 2 (CNN-LSTM) | MIMIC II | PPG | **3.99** | **6.40** | 2.65 | **4.71** |

Note: the maximum and minimum values of MAE and STD were in bold.

We further assessed the performance of model 2, the best method we proposed, based on Association for the Advancement of the Medical Instrumentation (AAMI) standard[44] and British Hypertension Society (BHS) standard[45] (Table 5). According to the AAMI standard, it requires the values of mean error (ME) and STD tested on more than 85 subjects lower than 5mmHg and 8mmHg separately. However, the BHS standard divides the performance of BP measuring devices into three grades based on the cumulative frequency percentage of errors (details were showed in Table 5). As a result, our proposed model 2 satisfied the AAMI standard and obtained grade A for SBP and DBP estimation according to the BHS standard.

**Table 5.**
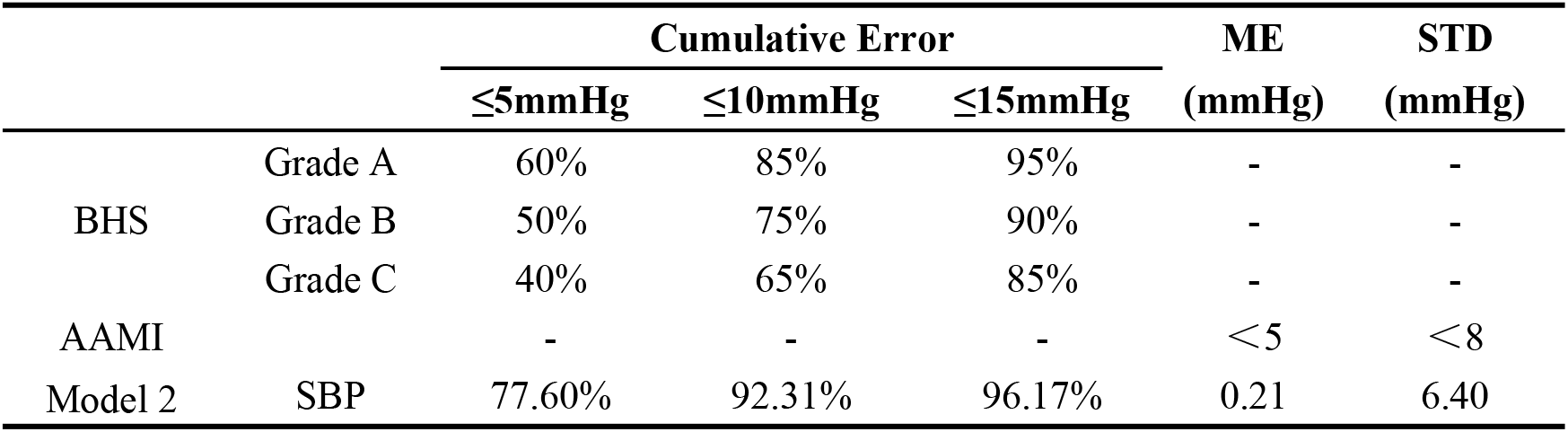
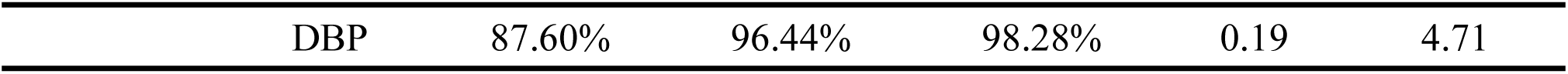
Performance evaluation based on AAMI and BHS standards.

The distributions of SBP and DBP estimation error from the model 2 were presented in Figure 8. Both of the distributions were similar to the normal distribution and distributed around zero, while, the estimation errors for DBP have a lower variation compared with the SBP, indicating that the less variances in the target data, the smaller the prediction error.

**Figure 8.**
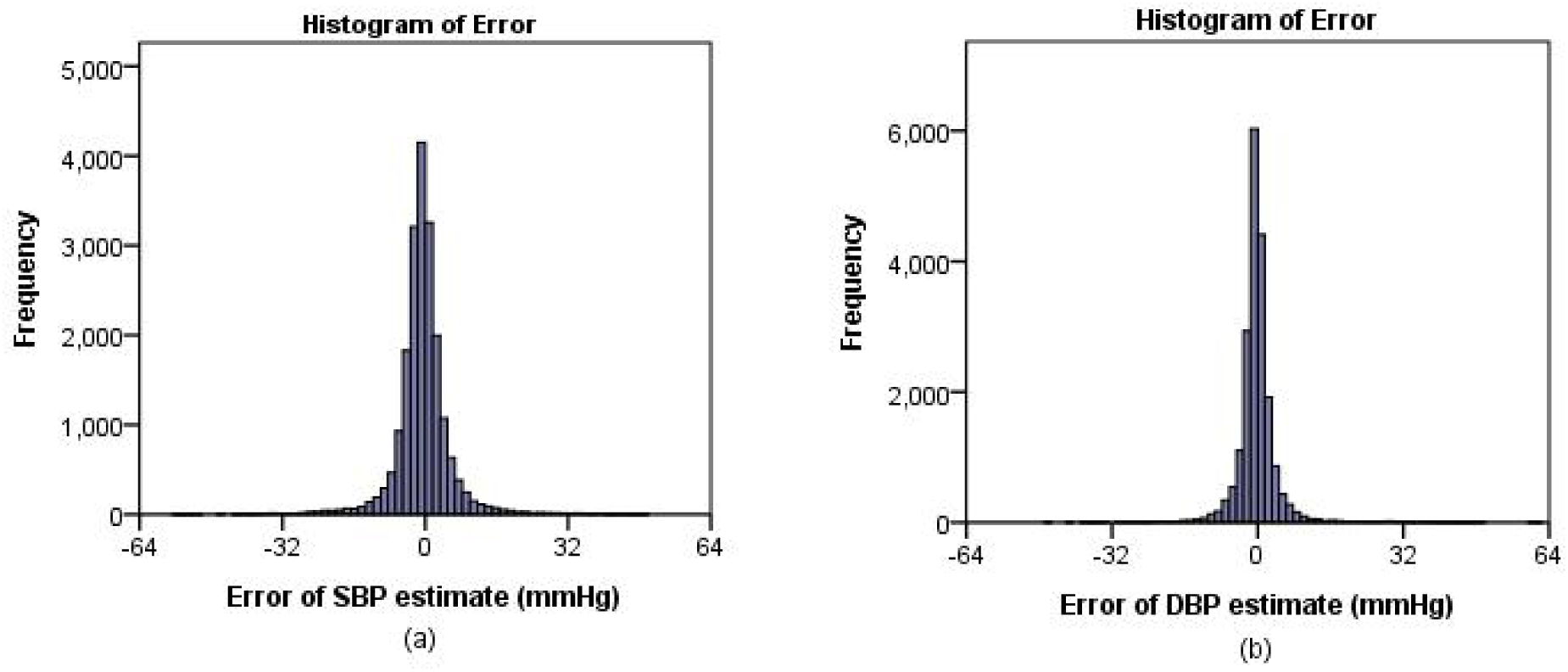
Error histogram from the model 2. (a) SBP, (b) DBP.

The spearman correlation coefficient between target and estimated values for SBPs and DBPs were 0.86 and 0.82 (Figure 9 a and b), respectively, indicating that there is a high correspondence between true and predicted BP. In addition, there was no significant difference in the distribution of the target and predicted values, although there were some outliers in the data (Figure 9 c and d). It means that our proposed model can also achieve satisfactory accuracy when predicting extreme BP values.

**Figure 9.**
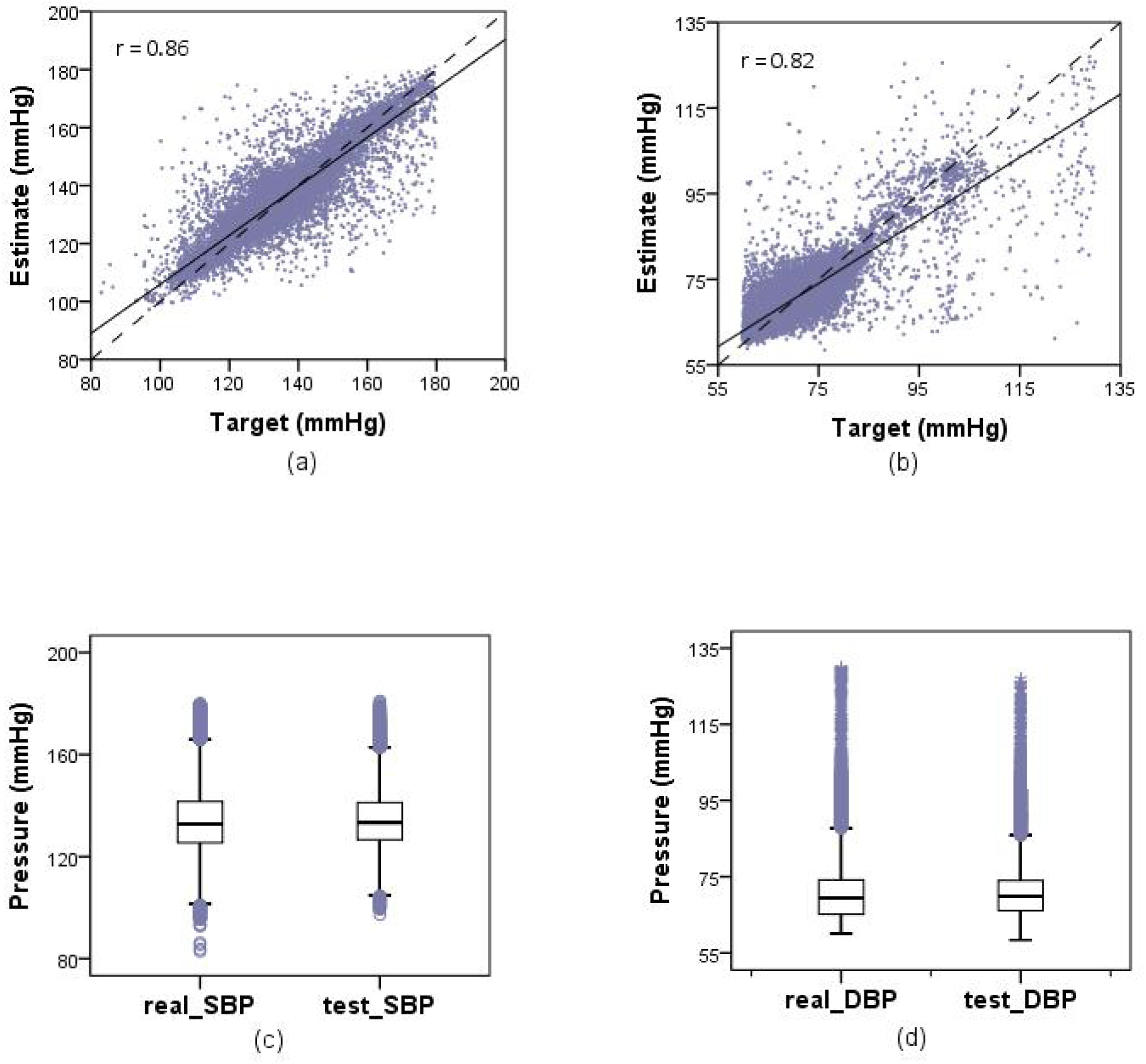
Regression plot and the box plot for SBP and DBP based on the estimation results from model 2. (a) and (b) represented the regression plot for SBP and DBP respectively, (c) and (d) represented the box plot for SBP and DBP respectively.

Moreover, we performed consistency evaluation using the Bland-Altman plots of SBP and DBP from model 2 (Figure 10). The area between two dashed lines represents that there is 95% of the difference between target and estimated values of SBP or DBP falling in [Mean - 1.96SD, Mean + 1.96SD], which is known as the 95% limits of agreement (95% LoA). Here, the 95% LoAs for SBP and DBP were [− 12.76, 10.76] and [− 9.80, 9.80], separately. As shown in figure 10, most of the errors fall within the ranges, thus, there is good consistency between the target and estimated values.

**Figure 10.**
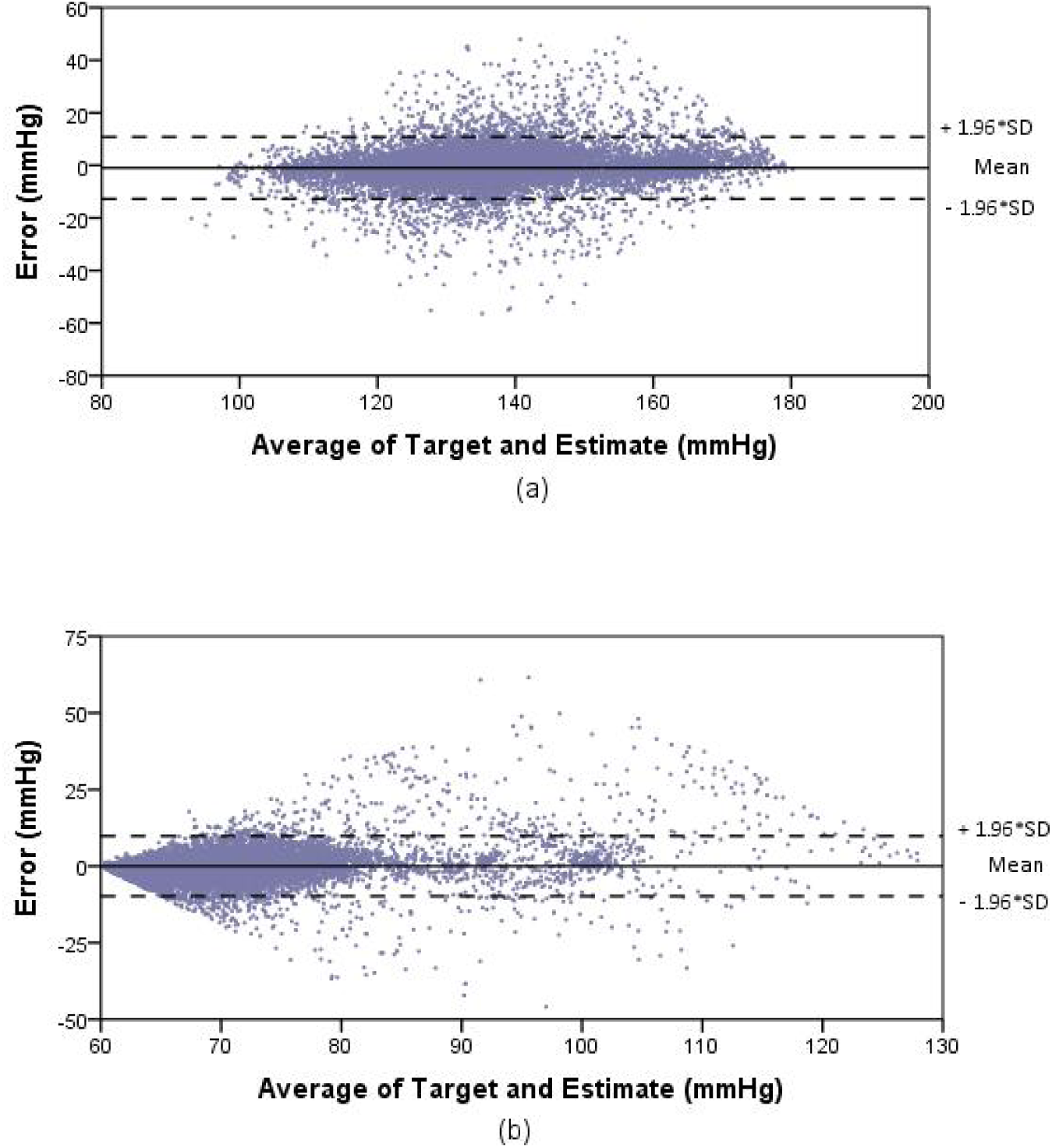
Bland-Altman plot of model 2. (a) SBP, (b) DBP.

## 5. Discussion

The subjects, quality of signals, data volume, and algorithms may influence the results of BP evaluation. In order to increase the comparability of the models, we chose to compare with other BP estimation methods based on datasets similar to ours in this paper and most of them came from the same batch of data initially,which became cleaner after rough preprocessing[35]. For instance, Sharifi, et al. [43] proposed a novel dynamical method by using PTT and PPG intensity ratio (PIR) coming from the preprocessed dataset for continuous BP estimation. Li, et al. [2] employed a deep learning model which had a similar framework with model 1 proposed in this work and adopted the same batch of data with our works, based on the ECG and PPG signals for BP estimation. Kachuee, et al. [26] presented a regression algorithm based on PAT and informative features from the vital signals in dataset MIMIC II for the continuous and cuff-less BP estimation. Such comparison of the performance of different models in predicting BP values based on similar dataset is hard to see in previous similar studies. The result shows that the estimation error of the deep neural network models generally lower than the traditional machine learning methods. As for the reason why the STD of SBP in ‘BiLSTM-4L’[2] is so high, it may be caused by the huge variation of SBP in the database. It can be seen that the fluctuation range of SBP and DBP in the training dataset has great influence on the results.

In addition, we compared the model 1 with the algorithm of [2] to investigate the influence of different input signals on the performance of results with similar frameworks (Table 2). The outcome indicates that the performance of models based on the unique PPG signals can achieve a comparable and even better effect as the models based on the ECG and PPG signals, which suggests that the unique PPG signals are also promising for estimating the continuous BP values. Models based on the unique PPG signals can be more conveniently applied to daily monitoring of continuous BP values.

BiLSTM and CNNs are commonly used for automatic feature extraction in the latest studies. In order to compare the pros and cons of the two, we built two models based on them respectively, namely model 1 and model 2. Finally, we found that the result of BP estimation model based on CNNs performed better, indicating that CNNs might be slightly better than BiLSTM in extracting PPG features. The reason behind this, presumably, is that CNNs are good at learning the morphological features of signals from the spatial dimension, which helps to discover the local characteristics of signals and preserve more details[46]. Of course, it still needs more studies to verify.

However, there are also some limitations in our study. For example, our training samples were all from patients in the intensive care unit, hence, our algorithm had not been verified on other populations, so that lacking the evidences to evaluate the generalization ability of the models. Another important point is that the quality of the original signals in the MIMIC database was uneven, so the performance of the models depended heavily on the effect of preprocessing. It may have better results if the stable and continuous high-quality signals can be obtained.

## 6. Conclusion and future work

In this paper, we proposed two multistage models based on deep neural network for estimating cuff-less continuous blood pressure using the unique PPG signals. Both models contained three parts, and the only difference between them was part one. The first part of model 1 adopted BiLSTM, while the model 2 adopted CNN. Although both of them had shown good prediction performance, the model based on CNN was slightly better. For the similar dataset, the performance of our proposed models were better than other models[2, 26, 43] which required manual feature extraction. For the similar framework, the model 1 based on the unique PPG signals we proposed performed better than the model based on the ECG and PPG signals of [2]. Moreover, our proposed models had absolute advantages in automatic feature extraction, which is able to achieve the continuous BP measurement. As the best model we proposed, the model 2 satisfied the AAMI standard and obtained grade A for SBP and DBP estimation according to the BHS standard. What’s more, it could be seen that the model had good fitting ability from the regression plots and the box plots for SBP and DBP. And the Bland-Altman plot showed that there were most errors fall within the 95% LoA, which indicated the good consistency between the target and estimated values. Nonetheless, all subjects of our proposed models came from the intensive care unit, the PPG signals of which were inferior in quality and lacks representativeness. Therefore, we must carry out complicated preprocessing in order to obtain high-quality PPG signals. In future works, we will try to improve the robustness and generalization of the model by training in larger number and wider variety of subjects with stable and high-quality PPG signals collected by more advanced equipments.

## Data Availability

https://www.kaggle.com/mkachuee/BloodPressureDataset

